# Hydroxychloroquine application is associated with a decreased mortality in critically ill patients with COVID-19

**DOI:** 10.1101/2020.04.27.20073379

**Authors:** Bo Yu, Chenze Li, Peng Chen, Ning Zhou, Luyun Wang, Jia Li, Hualiang Jiang, Dao Wen Wang

**Affiliations:** Division of Cardiology, Department of Internal Medicine and Hubei Key Laboratory of Genetics and Molecular Mechanisms of Cardiological Disorders, Tongji Hospital, Tongji Medical College, Huazhong University of Science and Technology, Wuhan 430030, China; State Key Laboratory of Drug Research, Shanghai Institute of Materia Medica, Chinese Academy of Sciences, 555 Zu Chong Zhi Road, Shanghai 201203, China; Shanghai Institute for Advanced Immunochemical Studies and School of Life Science and Technology, Shanghai Tech University, Shanghai 201210, China

## Abstract

**Importance:** Coronavirus disease 2019 (COVID-19) is a pandemic with no specific drugs and high mortality. The most urgent thing is to find effective treatments.

**Objective:** To determine whether hydroxychloroquine application may be associated with a decreased risk of death in critically ill COVID-19 patients and what is potential mechanism.

**Design, Setting and Patients:** This retrospective study included all 568 critically ill COVID-19 patients who were confirmed by pathogen laboratory tests despite antiviral treatment and had severe acute respiratory distress syndrome, PAO2/FIO2 <300 with need of mechanical ventilation in Tongji Hospital, Wuhan, between February 1 of 2020 to April 8 of 2020. All 568 patients received comparable basic treatments including antiviral drugs and antibiotics, and 48 of them additionally received oral hydroxychloroquine (HCQ) treatment (200 mg twice a day for 7-10 days). Primary endpoint is mortality of patients, and inflammatory cytokines levels were compared between hydroxychloroquine and non-hydroxychloroquine (NHCQ) treatments.

**MAIN OUTCOMES AND MEASURES:** In-hospital death and hospital stay time (day) were obtained, level of inflammatory cytokine (IL-6) was measured and compared between HCQ and NHCQ treatments.

**RESULTS:** The median age of 568 critically ill patients is 68 (57, 76) years old with 37.0% being female. Mortalities are 18.8% (9/48) in HCQ group and 45.8% (238/520) in NHCQ group (p<0.001). The time of hospital stay before patient death is 15 (10-21) days and 8 (4 - 14) days for the HCQ and NHCQ groups, respectively (p<0.05). The level of inflammatory cytokine IL-6 was significantly lowered from 22.2 (8.3-118.9) pg/mL at the beginning of the treatment to 5.2 (3.0-23.4) pg/ml (p<0.05) at the end of the treatment in the HCQ group but there is no change in the NHCQ group.

**CONCLUSIONS AND RELEVANCE:** Hydroxychloroquine treatment is significantly associated with a decreased mortality in critically ill patients with COVID-19 through attenuation of inflammatory cytokine storm. Therefore, hydroxychloroquine should be prescribed for treatment of critically ill COVID-19 patients to save lives.

**Key Points:** *Question:* Could hydroxychloroquine administration be beneficial in the treatment of critically ill patients with COVID-19?

*Findings:* In this retrospective study, a total of 568 critically ill patients with COVID-19 all received basic therapy and additionally 48 of them received hydroxychloroquine for 7–10 days (200 mg twice per day). Hydroxychloroquine treatment is significantly associated with a decreased mortality in critically ill COVID-19 patients and attenuated inflammatory cytokine IL-6 level.

*Meaning:* Our data suggest that hydroxychloroquine could be used to treat critically ill patients with COVID-19 which may save a lot of lives.

## Introduction

Since the breakout of infection by a novel severe acute respiratory syndrome coronavirus 2 (SARS-CoV-2) at the end of 2019 in Wuhan, China^1^, the Coronavirus Disease 2019 (COVID-19) has become a worldwide pandemic, with more than 1,918,432 infections and 123,460 death as reported on April 14 of 2020.

To combat this awful epidemic and help frontline physicians to treat patients and save lives, the World Health Organization (WHO), China, and many other countries have issued preliminary guidance on screening and diagnosis of infections in populations as well as managements of infection control. However, because of the lack of effective remedy for COVID-19, the recommended treatment for acutely ill patients are severely limited, with options for compassionate managements that include the use of Chinese herbs^2^. Although these strategies help many patients, especially in China, severe and especially critically ill patients are with high risk of death. Recent clinical trials on antiviral drugs such as Arbidol have not shown significant therapeutic benefit and the effect of Remdesivir still needs more evidence^3^. Recently clinical observation suggest that Tocilizumab against interleukin 6 (IL-6), a major proinflammatory cytokine, to calm down the cytokine storm caused by SARS-CoV-2 infection, could lead to therapeutic effects in portion of patients^4^. More recently, plasma of convalescent patients was used in Wuhan to manage refractory patients successfully but the scope of this treatment is greatly constrained due to the limited supply of effective plasma.

Chloroquine and hydroxychloroquine are two classic drugs^5^, which were originally used to treat malaria. In recent years, repurpose of existing drugs such as chloroquine class of drugs for other diseases has received increasingly growing attention^5,6^. Chloroquine and hydroxychloroquine have been used to treat rheumatological and immunological diseases because their inhibition of immunity and proinflammatory cytokines. Chloroquine and hydroxychloroquine also have antimicrobial effects and were used to treat infections by bacteria and virus. Recently, Liu et al. suggested that hydroxychloroquine is effective in inhibiting SARS-CoV-2 infection cell-based assays^7^ and Gautret et al demonstrated hydroxychloroquine treatment is significantly associated with reduction and disappearance of viral load in COVID-19 patients in a small size sample^8^, which instigated new hope for an effective treatment for COVID-19.

The key to reduce mortality of COVID-19, however, is to cure critically ill patients. This study demonstrates that hydroxychloroquine application, a less toxic derivative of chloroquine, is significantly associated with a decreased mortality in critically ill patients with COVID-19 by attenuating the inflammatory cytokine storm.

## Methods

### Study design

This investigation is a retrospective study involving critically ill patients with COVID-19 in Tongji Hospital, Wuhan, China. These patients were diagnosed according to the WHO interim guideline and the Clinical Guideline for COVID-19 Diagnosis and Treatment published by the National Health Commission of China between February 1 of 2020 and April 8 of 2020. This study has been approved by the institutional review board of Tongji Hospital (IRBID: TJ-C20200113). The written informed consent has been waived by the Ethics Committee because of the retrospective and anonymous nature of the data.

### Patients’ information

All patients have medical history and imaging characteristics of COVID-19 and have been confirmed with SARS-CoV-2 infection by laboratory tests. In addition, all patients were confirmed by chest computed tomography (CT) and SARS-CoV-2 pathogenic test. The inclusion for critically ill patients has to meet one of the following criteria: 1) patients had respiratory failure and needed mechanical ventilation; 2) patients had septic shock during hospitalization; 3) patients with other organ failures that required monitoring and treatment by intensive care unit. In this study, we included all critically ill patients with hospitalization during the epidemic period from February 1 of 2020 to April 8 of 2020.

### Data collection of patients

Health care data of COVID-19 patients, including medical history, chest CT, and laboratory tests, in-hospital therapies, and clinical deposits (death or cured discharge), were extracted by data coordinators through the electronic medical records. Laboratory test results included inflammatory cytokines and counts of white blood cells in peripheral blood.

### Statistical analysis

Continuous values were expressed as medians and interquartile range (Q1-Q3) and categorical variables as counts and percentages. The comparisons of continuous values between groups were performed with Wilcoxon rank-sum tests. For paired continuous variables, paired sample Wilcoxon rank test was used. Categorical variables were compared using Chi-square test. Survival curves was described by Kaplan-Meier method and compared with the log-rank test. Cox regression were performed to determine hazard ratios (HRs) and 95% confidence intervals (CIs). In the mulitivariable model, we adjusted for age, sex, history of hypertension, diabetes mellitus, coronary heart disease, and COPD, and oxygen saturation, as well as baseline treatment drugs. All comparisons were two-tailed, and p < 0.05 was considered as statistically significant. All statistical analyses were performed with R packages (version 3.1.4, Vienna, Austria).

## Results

### Study patients

From February 1 of 2020, through April 8 of 2020, a total 568 critically ill COVID-19 patients were admitted to Tongji Hospital, 358 males and 210 females. Median ages were 68 years old; 48 patients received hydroxychloroquine treatments (HCQ) (oral 200 mg twice per day for 7-10 days) and remaining 520 receiving basic treatments (non-hydroxychloroquine treatments, NHQC). Between these two groups, baseline characteristics including age, genders, original comorbidities, as well as severity of disease were not different (table 1).

**Table 1.** Baseline characteristics of critically ill COVID-19 patients

|  | All patients<br>(n = 568) | HCQ<br>(n = 48) | Non-HCQ<br>(n =520) | *P* |
| --- | --- | --- | --- | --- |
| Age, years | 68 (57-76) | 68 (60-75) | 68 (57-77) | 0.933 |
| Age range, years |  |  |  | 0.444 |
| ≤60 (%) | 157 (27.6) | 11 (22.9) | 146 (28.1) |  |
| >60 (%) | 411 (72.4) | 37 (77.1) | 374 (71.9) |  |
| Gender, male (%) | 358 (63.0) | 32 (66.7) | 326 (62.7) | 0.585 |
| Original comorbidities |  |  |  |  |
| Hypertension (%) | 252 (44.4) | 23 (47.9) | 229 (44.0) | 0.605 |
| Coronary heart disease (%) | 59 (10.4) | 2 (4.2) | 57 (11.0) | 0.213 |
| COPD (%) | 16 (2.8) | 0 (0) | 16 (3.1) | 0.384 |
| Diabetes (%) | 97 (17.1) | 12 (25.0) | 85 (16.3) | 0.127 |
| SpO<sub>2</sub> on admission (%) | 96 (90-98) | 94.5 (90-96) | 96 (90-98) | 0.216 |
| Oxygen therapy, n (%) | 547 (96.3) | 47 (97.9) | 500 (96.2) | 0.714 |
| Mechanical ventilation, n (%) | 349 (61.4) | 28 (58.3) | 321 (61.7) | 0.644 |

Abbreviations: HCQ, hydroxychloroquine; COPD, chronic obstructive pulmonary disease; SpO<sub>2</sub>, percutaneous oxygen saturation.
The percentage represented the frequency divided by the total cohort size (n=568).
Mechanical ventilation contained Non-invasive ventilation and Invasive ventilation
Data were presented as medians and interquartile range (Q1-Q3).
HCQ, Hydroxychloroquine treatment; NHCQ, non-Hydroxychloroquine treatment.

### Study outcomes

In total of 568 patients, 247 patients died (mortality was 43.5%). Nine out of 48 HCQ patients (18.8%) died, while in NHCQ group, 238/520 (45.8%) patients died (*p*<0.001). Furthermore, average hospital stay time are 32 (26-41) days in HCQ group and 30 (18-40) days in NHCQ-treated patients (*p*=0.098), but hospital stay time before death from admission was longer in HCQ patients than NHCQ subjects [15 (10-21) vs. 8 (4-14)] days (p=0.021) (Table 2, and Figure 1A).

**Figure 1.**
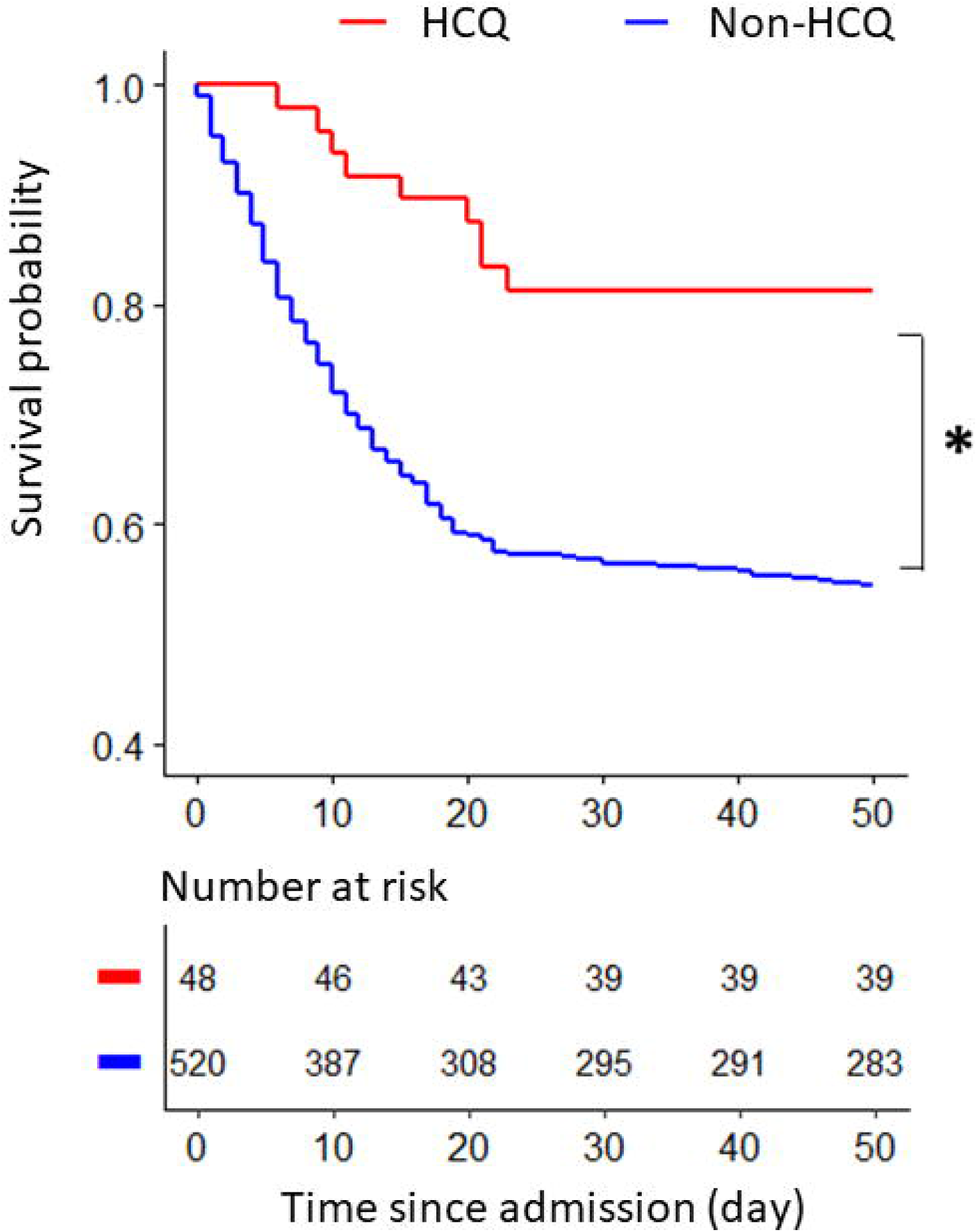
Kaplan-Meier curve of COVID-19 patients treated with or without hydroxychloroquine. Hydroxychloroquine treatment is significantly associated with a decreased mortality in critically ill COVID-19 patients compared with those without hydroxychloroquine treatment (*P<0.001). HCQ, hydroxychloroquine; NHCQ, non-hydroxychloroquine.

**Table 2.**
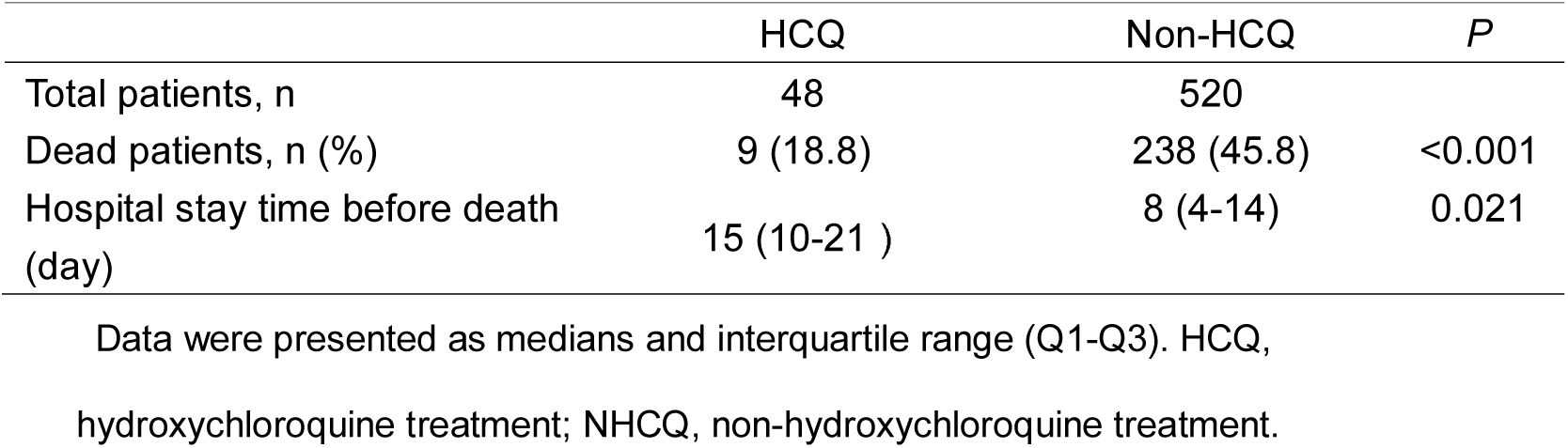
Comparison of clinical outcomes between HCQ treated and non HCQ treated patients.

|  | HCQ | Non-HCQ | *P* |
| --- | --- | --- | --- |
| Total patients, n | 48 | 520 |  |
| Dead patients, n (%) | 9 (18.8) | 238 (45.8) | <0.001 |
| Hospital stay time before death (day) | 15 (10-21 ) | 8 (4-14) | 0.021 |

Data were presented as medians and interquartile range (Q1-Q3). HCQ, hydroxychloroquine treatment; NHCQ, non-hydroxychloroquine treatment.

The baseline treatments were comparable for these two groups, including application of antiviral drugs (Lopinavir and Ritonavir, Entecavir hydrate, or Ribavirin) with 41.7% and 44.4% patients in HCQ and NHCQ, respectively, (p=0.71); intravenous immunoglobulin in 52.1% in HCQ and 47.1% patients in NHCQ, respectively (p=0.51); immunoenhancer in 16.7% in HCQ and 17.3% patients in NHCQ, respectively (p=0.91), but antibiotics in 77.1% in HCQ and 89.4% patients in NHCQ, respectively (p=0.01); but interferon application 0% in HCQ and 10.4% patients in NCHQ (p=0.01).

In order to eliminate the influence of confounding factors, we performed Cox regression analysis for age, sex, history of hypertension, diabetes mellitus, coronary heart disease, and COPD, and oxygen saturation, and results showed that the use of hydroxychloroquine was associated with a significantly decreased mortality risk (unadjusted HR: 0.33; 95% CI: 0.17-0.64; p=0.001; adjusted HR: 0.32; 95% CI: 0.16-0.62; p<0.001), Furthermore, we adjusted effect of various baseline treatment drugs and results still showed that the use of hydroxychloroquine was associated with a significantly decreased mortality risk (unadjusted HR: 0.33; 95% CI: 0.17-0.64; p=0.001; adjusted HR: 0.33 (0.17-0.65, p=0.001), which further supports that the critically ill patients can obtain substantial benefit from hydroxychloroquine treatment.

### Effect of hydroxychloroquine on inflammatory cytokines

Laboratory tests showed that in HCQ group, IL-6 level in plasma was 5.2 (3.0-23.4) pg/mL after HCQ application, which significant lower than the IL-6 level before HCQ application (22.2 [8.3-118.9] pg/mL, p=0.002), but there is little change in NHCQ patients (21.3 [8.8-62.8] pg/mL vs. 20.2 [6.1-94.4] pg/mL, p 0.05] (Figure 2). Furthermore, we analyzed the change of IL-6 level during the hospitalization period and found that IL-6 level was lowered after HCQ application and kept the lower level stable during therapy period. However, interestingly, after hydroxychloroquine treatment stopped, inflammatory mediator IL-6 level went up to control level (Figure 3).

**Figure 2.**
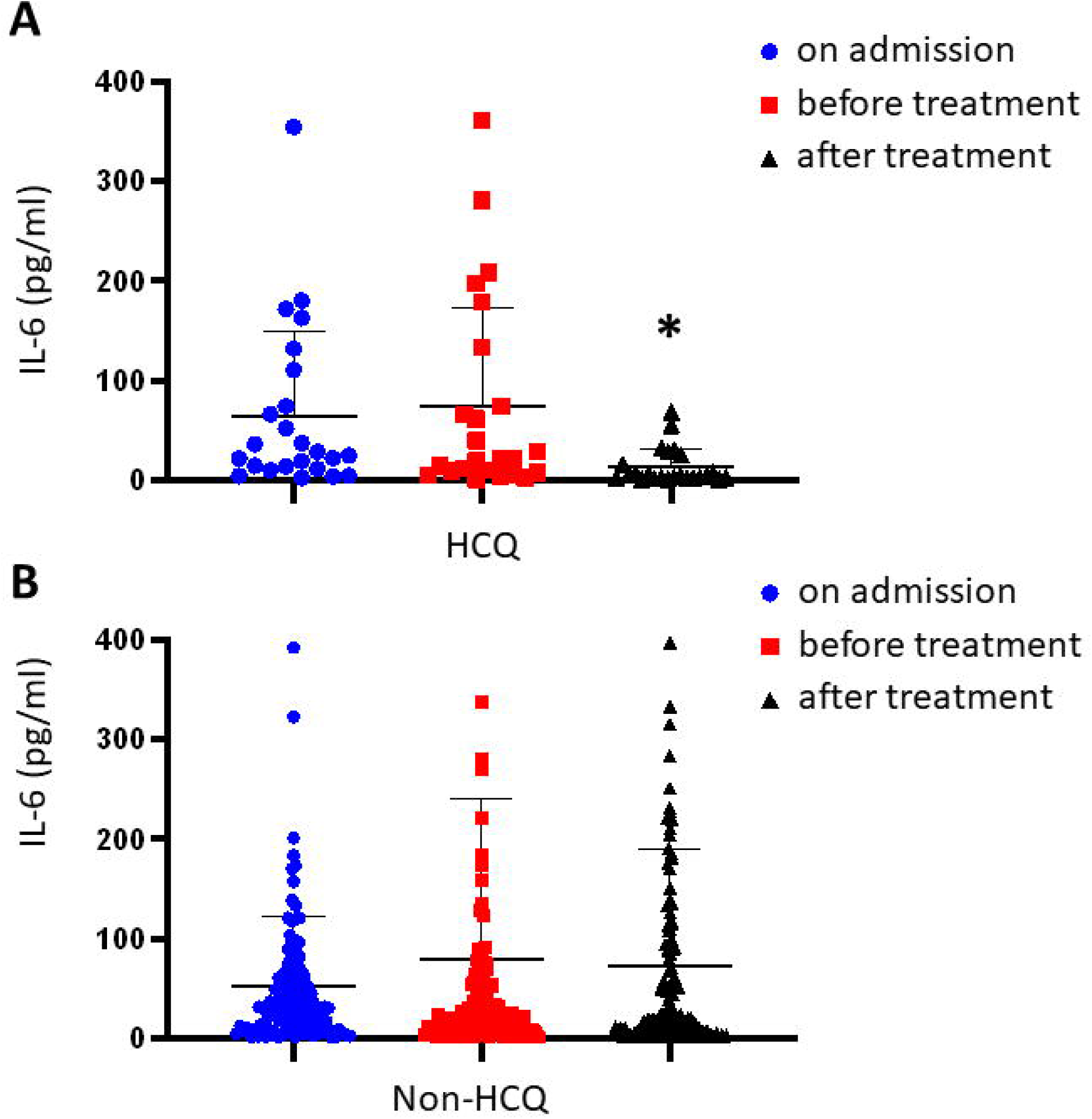
Effects of hydroxychloroquine treatment on plasma level of IL-6. Hydroxychloroquine treatment is associated with a decreased level of IL-6 in about 5 (4–8) days, *P<0.05 (A). The level of IL-6 did not change in non-hydroxychloroquine treatment group at the same observation time (B). HCQ, hydroxychloroquine; NHCQ, non-hydroxychloroquine.

**Figure 3.**
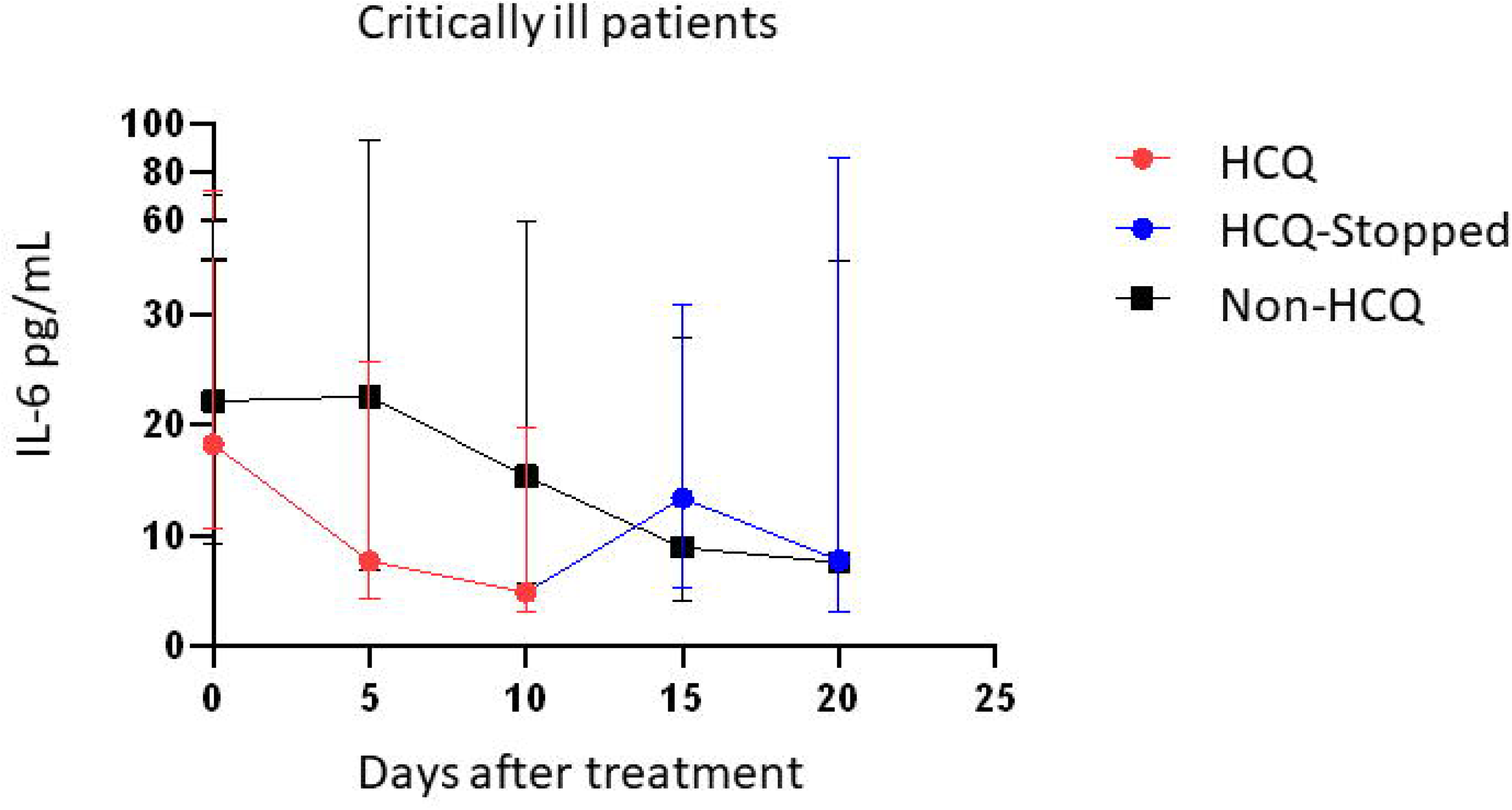
Continuous curve of plasma levels of IL-6 showing that the consistency of hydroxychloroquine treatment period with lowering IL-6 level.

## Discussion

This retrospective study found that hydroxychloroquine application is significantly associated with a decreased mortality in critically ill patients with COVID-19 and lower level of IL-6, one of most inflammatory cytokines. We also found that the length of hospital stay in the hydroxychloroquine-treated non-survivors is longer than in non-hydroxychloroquine-treated non-survivors, suggesting that hydroxychloroquine could prolong the lifetime of critically ill patients even if their lives were eventually lost, which further supports that hydroxychloroquine has therapeutic effects for critically ill patients of COVID-19. Importantly, we found that the role that hydroxychloroquine application associated with a decreased plasma IL-6 level is highly consistent with the duration of its administration, and once the drug stops, plasma IL-6 level rises up to control level.

We have noted that hydroxychloroquine has in vitro antiviral effects on several viruses, including SARS, and SARS-CoV-2^7–10^, which may contribute directly to its therapeutic effect to COVID-19 patients. However, the anti-SARS-CoV2 action of chloroquine does not seems to account for the significant efficiency to critically ill patients in our cohort.

COVID-19 patients displayed obvious immunity disorders, showing marked reduction in lymphocyte numbers in peripheral blood and lymphatic tissues, but huge amount of lymphocyte infiltration in lungs as well as in other critical organs such as heart. Many patients showed symptoms of cytokine storm with markedly-elevated levels of the proinflammatory cytokine IL-6, suggesting overactivated immune responses^11^. Anti-IL-6 antibody, Tocilizumab, has been been shown to be an effective treatment option for COVID-19 patients with a risk of cytokine storms.^12^ In this study, we demonstrate that hydroxychloroquine can mimic the effect of anti-IL-6 antibody by observing decreased levels of Il-6 in the critically ill COVID-19 patients after hydroxychloroquine application. In addition, hydroxychloroquine can modulate human inflammatory macrophage polarization via downregulating M1 but upregulating M2 macrophages^13,14^, and inhibit proinflammatory cytokines through inhibition of lysosomal-autophagy pathways^14^ and formation of double membrane vesicles ^6,15^, a process required for genome replication by the SARS Coronavirus Replication Complex^16^. Thus, its anti-inflammatory action of hydroxychloroquine in combination of its inhibitory of viral replication may contribute greatly to its therapeutic effects on critically ill COVID-19 patients. In addition, tissue distribution of hydroxychloroquine is unique and favors therapy of pneumonia because it has much higher concentrations in lungs^17^. Importantly, the chloroquine class of drugs have been showed to repress inflammation with great synergy with low concentrations of glucocorticoids, one of the most potent classes of anti-inflammatory drugs, pointing to possibility of combined therapy of hydroxychloroquine with low dose of glucocorticoids^14^.

Additionally, a decreased plasma IL-6 level after hydroxychloroquine use is highly consistent with its application period and after stopped its role disappear and IL-6 level increases to the level of non-hydroxychloroquine patients. This finding indicates that we need to adjust our treatment regimen, that is, to keep use hydroxychloroquine until patients are completely cured.

In summary, this retrospective study demonstrates that hydroxychloroquine application is associated with a decreased risk of death in critically ill COVID-19 patients without obvious toxicity and its mechanisms of action is probably mediated through its inhibition of inflammatory cytokine storm on top of its ability in inhibiting viral replication. Therefore, hydroxychloroquine should be considered as a primary option to apply to treat critically ill COVID-19 patients, which could save lives during the current COVID-19 pandemic despite that randomized-double blind control study is needed to provide stronger evidence. Although this retrospective study was performed with the critically ill COVID-19 patients, hydroxychloroquine should also be an option for early stage of patients because the safety records and their long history of use in treating malaria infections.

## Data Availability

The data that support the findings of this study are available from the
corresponding author on reasonable request.

## Author announcements

1. Conflicts of interest: The authors have declared that no competing interests exist.
2. Its content, figures, and tables have not been published or submitted elsewhere in print or electronic format and will not be submitted elsewhere during the period of review by ***SCIENCE CHINA Life Sciences***.

## Acknowledgments

We thank all our colleagues from the Department of Internal Medicine, Tongji Hospital, as well as all the medical staffs fighting against COVID-19, for their tremendous efforts. We also thank Professor H. Eric Xu in Shanghai Institute of Materia Medica, Chinese Academy of Sciences for his great help in writing this paper.

